# From diagnosis to daily care: The impact of early informational support on the burden of family caregiving

**DOI:** 10.1101/2025.07.25.25332027

**Authors:** Tomoko Wakui, Tsuyoshi Okamura, Kae Ito, Takumi Hirata, Tomoyuki Yabuki

**Author notes:** **Corresponding Author Tomoko Wakui**, Tokyo Metropolitan Institute for Geriatrics and Gerontology.

## Abstract

People with dementia and their families receive a flood of wide-ranging information at diagnosis. However, it remains unclear which types of information are helpful to caregivers during the care period that follows. In collaboration with dementia medical centers and certified dementia support doctors, we distributed questionnaires to family caregivers of individuals diagnosed with dementia. A total of 159 self-administered questionnaires were retrieved. Caregiver burden was measured using the short version of the Japanese Zarit Caregiving Inventory (J-ZBI_8). Regression analyses were performed to examine how nine types of informational support provided at diagnosis were associated with caregiver burden while controlling for care recipient and caregiver characteristics and the caregiving context. Only dementia-related information and information from local medical institutions were significantly associated with a lower caregiver burden. These basic forms of information are fundamental yet crucial to enable family caregivers to navigate the subsequent stages of care.

**Clinical Trial Registration:** **Not applicable – this study did not involve a clinical trial.**

## Introduction

Receiving a diagnosis of Alzheimer’s disease or related dementia (ADRD) can be a highly emotional and difficult moment for families that results in both benefits and challenges for family caregivers.^1-5^ For some families, a diagnosis provides confirmation that previously concerning behaviors are indeed symptoms of ADRD^1,3^; for others, diagnosis may arrive as an unexpected and overwhelming disclosure^5^ with no clear solutions or information available.^4-7^ However, diagnosis also provides critical opportunities to access appropriate knowledge, caregiver training, and resources for managing daily care and participating in healthcare planning for the person with ADRD.^2,5,7^

Importantly, because effective treatments for ADRD remain limited, the way the diagnosis is communicated as well as the quality of informational and emotional support offered at this early stage can strongly shape both individuals’ and families’ coping responses. Many family members report receiving insufficient or inconsistent information regarding the condition and available support services,^4,8,9^ and the level and quality of information may vary considerably by provider and setting.^9,10^ Adequate and timely information at this stage is therefore critical to help families better understand the diagnosis,^10^ reduce confusion and anxiety,^11^ and facilitate earlier access to appropriate services,^9^ which may in turn help ease the caregiving burden as daily care needs become more complex.

The caregiving burden has been widely studied, and various contributing factors have been described, such as in Pearlin’s stress process model. The burden of caring for people with ADRD arises from multiple dimensions and includes stressors related to the care recipient’s dependence in both basic and instrumental activities of daily living (ADL) and behavioral and psychological symptoms of dementia (BPSD).^12^ In addition to these primary stressors, role-related demands, such as conflicts between caregiving and work, raising children and caregiving, and/or conflicts with relatives, are factors.^12^ However, the way the experience of receiving a diagnosis fits into this stress framework has not been fully examined.

Although various stressors contribute to the burden of caregiving, some factors, particularly information support, may help to alleviate this strain for family caregivers Informational support, which is recognized as an important dimension of social support, works alongside instrumental support (i.e., direct help with care tasks) and emotional support (i.e., empathy, understanding, and listening) to enable family members to cope with daily caregiving, reduce caregiver burden,^11,13^ and support caregivers’ psychological well-being and sustained engagement in caregiving roles.^1,3,5,14^ Despite its recognized importance, there is limited quantitative evidence on how specific types of informational support provided at diagnosis affect the subsequent caregiving experience and burden.

Few studies have focused on the specific types of information discussed or provided. Prior research on the needs of family caregivers of individuals with ADRD indicates that caregivers often prioritize information related to diagnosis, treatment, and legal or financial matters, especially health plan coverage, over general disease knowledge.^9,15^ Studies also suggest that these needs may vary by caregiver characteristics; for example, experienced caregivers and female caregivers tend to report stronger needs for legal, financial, and emotional support.^15^ In addition, qualitative work has shown that families need clear information about the progression of dementia toward the end of life, decision-making roles, and sustaining connections.^16^ Informational support may address both disease-specific issues and the broader context of caregiving.

In Japan, individuals living with dementia and their families rely on a dual support system of national medical insurance and long-term care insurance (LTCI) programs. Ideally, medical needs, such as diagnostic procedures, treatment of comorbid conditions, and symptom management, are addressed through medical insurance, whereas daily care needs, including personal care, home-based services, and institutional care, are covered under the LTCI scheme. In practice, however, ensuring a smooth transition from medical diagnosis to daily caregiving remains challenging.^17-19^ Gaps in coordination between the medical and long-term care sectors combined with limited informational support for families can delay timely access to appropriate services^8,20^ and increase caregiver burden.^4,7,16^

In summary, receiving a dementia diagnosis is a pivotal moment that can profoundly shape family caregivers’ experiences. Although various factors that influence the subsequent caregiving burden have been identified, few studies have quantitatively examined how information provided at diagnosis affects caregivers’ long-term stress and preparedness. While informational support is known to ease caregiving, existing evidence, which is largely qualitative, offers limited insight into which types of information are most useful at this stage. Clarifying this issue may strengthen the link between diagnosis and daily care and help families navigate transitions more effectively.

This study examines the effects of informational support provided to family members at the time of a dementia diagnosis via a caregiver survey and multivariate analysis that accounts for known factors associated with the burden of caregiving. Specifically, it addresses three key questions:

1. What types of information do families receive at the time of diagnosis?
2. What types of informational support are associated with a lower overall caregiving burden?
3. Are particular types of information linked to reduced personal strain and role strain, the two subcomponents of the caregiving burden?

By addressing these questions, this study seeks to clarify how hospital-based informational support can bridge fragmented care systems and reduce caregiver burden by helping families navigate complex care arrangements. The findings aim to inform the development of integrated, family-centered support pathways that promote continuity of care and sustained home caregiving.

## Methods

### Sampling

Sampling for this study was conducted in two stages. First, the researchers contacted all dementia-related medical centers across Japan (n =505) as well as local clinics affiliated with dementia support physicians in Northeast Japan (n=500) to request their cooperation. Dementia-related medical centers (*Ninchishō Shikkan Iryō Sentā*)^21^ are designated by each prefectural government in collaboration with the Ministry of Health, Labor and Welfare (MHLW) to serve as regional hubs for dementia care. These centers provide specialized outpatient diagnosis and treatment for individuals with suspected dementia and play a central role in coordinating care across hospitals, clinics, community support centers, and long-term care services. They also offer consultation and educational support to patients, family caregivers, and healthcare professionals and are typically located within general hospitals, psychiatric institutions, or university-affiliated facilities. In parallel, dementia support physicians^22^ are medical doctors who have completed a nationally accredited training program to increase their expertise in dementia care. These physicians act as liaisons between primary care providers and dementia specialists and support early diagnosis, care planning, and community-based collaboration. Their responsibilities include providing clinical guidance to other healthcare professionals, managing BPSD, and leading educational initiatives within their regions. Together, these two entities form the backbone of Japan’s regional dementia care infrastructure.

Each institution or clinic was asked to distribute 10 sets of questionnaires to family caregivers of patients diagnosed with dementia. Facilities that agreed to participate were mailed the questionnaire packages by the research team. The packages included a survey form, instructions, and a return envelope. The participating facilities distributed the questionnaires to the family members of patients who had been diagnosed with dementia.

In total, 78 dementia-related medical centers (15.0%) and 27 clinics affiliated with dementia support physicians (5.0%) agreed to collaborate, resulting in the distribution of 1,050 questionnaire sets between January and April 2025. Of these, 216 responses were returned by family caregivers, yielding a response rate of 20.6%.

### Measures

Caregiver burden was measured using the Japanese short version of the Zarit Caregiver Burden Interview (J-ZBI-8), a validated instrument that is widely used in dementia caregiving research in Japan. The J-ZBI-8 consists of 8 items rated on a 5-point Likert scale (0 = never to 4 = nearly always) and yields a total score that ranges from 0 to 32, with higher scores indicating greater perceived burden. In addition to the total burden score, two subscales were analyzed: *personal strain*, which reflects the emotional and physical stress experienced directly by the caregiver (e.g., fatigue, anxiety), and *role strain*, which captures stress related to disruptions in the caregiver’s social roles and responsibilities (e.g., conflicts with work or family life). The instrument has demonstrated good reliability and validity in Japanese caregiving populations.^23-25^

To examine the types of informational support provided at the time of diagnosis, we developed nine categories on the basis of previous research^9,15,16^ and clinical experience. The categories were refined through interdisciplinary discussions among the research team, which included professionals in psychiatry, public health, and social welfare. Additionally, volunteer family caregivers with experience supporting a relative through a dementia diagnosis were invited to review the questionnaire for content relevance and clarity.

Each type of informational support was assessed using binary (yes/no) response options. Caregivers were asked whether they had received information in each of the following nine domains: (1) basic information about dementia; (2) long-term care services and community-based integrated care programs; (3) legal support, including adult guardianship, abuse prevention, and independent living assistance; (4) financial support systems, including pensions, public assistance, and medical expense subsidies; (5) family caregiver support groups; (6) peer support meetings for persons living with dementia; (7) local medical institutions and clinics; (8) psychological support services for persons with dementia and their family members; and (9) employment support for caregivers, including caregiving leave and work-care balance programs. Each item was coded 1 for “received” and 0 for “not received.” These binary variables were used as individual predictors in the analysis to examine the associations between specific types of informational support and caregiver burden.

**Caregivers’ demographic and background characteristics,** including gender, age, employment status, financial status, subjective health, and coresident status with the care recipient, were collected through self-reports. For the care recipients, information was collected on age, gender, type of diagnosis, the presence of BPSD, and the number of years since the diagnosis of dementia to evaluate their overall care needs. BPSD were assessed using the Japanese short-form Dementia Behavior Disturbance Scale (DBD; 5 items).^26^

### Statistical Analysis

The analytic sample comprised 159 respondents who provided complete data on all variables included in the analysis, out of the 216 total respondents. Descriptive statistics were obtained to summarize the basic characteristics of participants and their family members living with dementia as well as the extent to which family caregivers received each type of informational support following the diagnosis. To examine the impact of informational support on the caregiver burden, multiple regression analyses were performed. First, a series of individual regression models were constructed to assess the associations between each of the nine types of informational support provided at the clinics and the overall caregiver burden while controlling for caregiver demographics (age, gender, employment status, and financial status) and caregiving-related factors, including the age and gender of the care recipient, the presence of BPSD, and time since diagnosis. A final regression model was subsequently constructed that included all nine informational support variables simultaneously to evaluate the unique effect of each type of support after adjusting for the presence of other supports and covariates.

Additionally, to explore whether different types of informational support were differentially associated with specific aspects of caregiver stress, we conducted separate regression analyses using the two subscales of the burden measure—personal strain and role strain—as dependent variables. These subscale analyses followed the same modeling procedures and covariates as the main models and aimed to clarify how distinct dimensions of the caregiver burden may be influenced by the nature of the information provided at diagnosis.

IBM SPSS Statistics version 29.0 was used for all the analyses, with statistical significance defined as p < .05.

### Ethical Considerations

This study was approved by the Tokyo Metropolitan Institute of Gerontology Institutional Review Board, no. R24-084. This study complied with the Declaration of Helsinki and its amendments or comparable ethical standards for conducting surveys. Participation by returning the completed questionnaire was considered to indicate consent. A checkbox was provided on the questionnaire for participants who wished to withdraw. Responses that were returned with this box unchecked were regarded as consent to participate in the study.

## Results

The majority of caregivers were women (66.7%), with an average age of 63.2 years (SD = 11.6). With respect to employment status, 56.0% were employed, 42.1% were unemployed, and 1.9% were currently on leave from work. In terms of perceived financial status, 5.7% rated themselves as very wealthy, 21.4% as wealthy, 40.9% as average, 23.9% as somewhat poor, and 8.2% as poor.

The care recipients were predominantly female (69.2%), with a mean age of 80.4 years (SD = 8.8). The average score on the 5-item Japanese version of the DBD-5 was 1.8 (SD = 1.4), indicating a mild level of BPSD. The mean time since diagnosis was 4.2 years (SD = 3.6). The mean caregiver burden, as measured by the Japanese short form of the J-ZBI_8, was 13.4 (SD = 8.2).

At the time of dementia diagnosis, caregivers reported receiving varying levels of informational support from hospital-based providers. The most frequently provided information concerned the medical condition itself, with 73.6% of caregivers reporting that they received information about dementia. Information related to long-term care services and community-based integrated care programs was received by 58.5% of the caregivers.

In contrast, support in more specialized or practical domains was less commonly reported. Only 22.6% of caregivers received information about legal support, including adult guardianship, abuse prevention, and independent living assistance, and 22.0% received information on financial support systems, such as pensions, public assistance, and medical expense subsidies. Psychological support services for persons with dementia and their family members were mentioned by 32.1% of the respondents, and 32.7% received information about family caregiver support groups. Peer support meetings for persons living with dementia were reported by 25.2% of caregivers, whereas 35.8% received information on local medical institutions and clinics. Notably, only 10.7% of the caregivers reported receiving information about employment support, including caregiving leave and work–care balance programs.

A series of regression analyses were conducted to examine the associations between different types of informational support received at the time of dementia diagnosis and subsequent caregiver burden. In Models 1–9, each type of support was entered separately. Dementia-related information was consistently associated with a significantly lower burden (B = –5.07, 95% CI = –7.81 to –2.38, p < .001), as was information about local medical institutions or clinics (B = –2.84, 95% CI = –5.44 to –0.25, p = .03). Information on family caregiver support groups showed a marginal association (B = –2.37, 95% CI = –5.00 to 0.27, p = .08), whereas other domains, including long-term care services and legal, financial, psychological, peer, and employment support, were not significantly associated with caregiver burden.

In Model 10, which included all nine types of support simultaneously, dementia-related information (B = –5.48, 95% CI = –8.46 to –2.51, p < .001) and local clinic information (B = – 3.12, 95% CI = –5.93 to –0.31, p = .03) remained significant, confirming their independent associations.

Further analysis of the subscales revealed that dementia-related information was significantly associated with lower personal strain (B = –3.16, p = .00) and role strain (B = – 2.35, p = .00). Local clinical information was also linked to reduced personal strain (B = –2.06, p = .03) but was not significantly linked to role strain (B = –1.06, p = .08). No other informational domains showed significant associations with either subscale.

## Discussion

This study shows that certain types of informational support provided at the time of dementia diagnosis have enduring effects that extend into later stages of care, especially by helping to reduce the burden experienced by family caregivers. These findings suggest that early-stage informational support not only is important for facilitating acceptance of the diagnosis and initiating service use^1,3^ but also plays a critical role in shaping long-term caregiving experiences.^4,7,9,16^

This study revealed a consistent association between receiving dementia-related information at the time of diagnosis and lower levels of subsequent caregiver burden, underscoring the protective role of basic disease education in the early stages of caregiving. Such information likely serves as a foundation for caregivers to develop an informed understanding of the condition,^3,14-16^ including typical symptoms, expected disease progression, and practical approaches to care. It can also reduce uncertainty, facilitate planning, and foster a greater sense of control over dementia-related behaviors and situations.^14^ This reflects the information family caregivers report wanting. Without such information and communication, receiving only the name of a diagnosis can be a particularly painful experience for families.^6,8,14^ Providing dementia-specific information that includes not only the name of the diagnosis but also typical symptoms, the expected disease progression, and practical approaches to care at the time of diagnosis should be considered key components of caregiver support strategies in the clinical setting.

This study also revealed that receiving information about local medical institutions at the time of diagnosis was independently associated with a lower caregiver burden. This finding highlights the importance of medical continuity in supporting families after diagnosis. Clear information about where and how to access ongoing care, such as memory clinics or community-based dementia specialists, can reduce caregivers’ anxiety and uncertainty about future care needs.^17^ This information offers not only a practical roadmap to services but also reassurance that professional guidance will remain available as symptoms progress.

From a care planning perspective, equipping caregivers with locally relevant and accessible medical contacts may serve as a key intervention point. These connections can prevent caregivers from navigating healthcare and social systems alone and contribute to more coordinated, community-based dementia care (e.g., as promoted in Japan’s regional comprehensive care system).^27,28^ However, not all local clinics have sufficient expertise in dementia care, leaving families to manage complex and evolving issues, including comorbidities and general health concerns, without adequate support. In this context, access to dementia-trained professionals, such as dementia support physicians, can be especially valuable. Even for nondementia-related concerns, providers who understand the caregiving context can improve communication, anticipate needs, and reduce caregiver stress.^8^ Strengthening these connections through timely, targeted information at diagnosis represents a practical strategy for building stable, integrated, and sustainable community-based dementia care.^4,7,9^

Unlike dementia-specific and local clinic information, other informational domains, such as long-term care services, legal support, financial assistance, and psychological services, did not show significant associations with caregiver burden in either the individual or the combined regression model. While these areas are undoubtedly important in the broader context of dementia care,^9,15,16^ their impact on caregiver burden may be limited during the initial stages following diagnosis. One possible explanation is that these forms of support may have lower perceived relevance or urgency at the diagnostic stage, when caregivers are focused primarily on understanding the condition and establishing basic care routines. Legal or financial procedures, as well as psychological support services, may be less immediate needs and therefore may be underutilized or overlooked by families in the early phases of caregiving.

Notably, information about family peer support groups demonstrated a marginal association with lower caregiver burden in the individual model, although the effect did not remain significant in the full model that included all informational domains. While peer support is often assumed to provide emotional relief through shared empathy,^29,30^ our findings suggest that caregivers may seek something more specific: practical, experience-based knowledge from other families that are navigating similar situations at the early stage of diagnosis. In this sense, peer groups may function as a complementary source of information^4^ to fill in gaps left by professional explanations, offer insights into real-world care decisions, and provide reassurance grounded in lived experience. These findings suggest that peer-based informational exchange may play a unique and nuanced role, not only emotionally but also as a practical coping tool at the beginning of care. Future research should explore how to formalize and increase the informational value of peer support programs provided after a dementia diagnosis to emphasize timely delivery during the early adjustment phase and ensure accessibility and credibility for caregivers as they begin navigating the dementia care process.

In summary, this study provides quantitative evidence that certain types of informational support at the time of dementia diagnosis—specifically, information about dementia and local medical institutions—are significantly associated with lower caregiver burden. The analysis also reveals that these types of information differentially reduce personal strain and role strain, the two key components of caregiver burden. These findings highlight the practical value of providing targeted, relevant information early in the care trajectory to support caregivers’ preparedness and reduce long-term stress. This study contributes new data-driven insights to inform the design of more effective, diagnosis-linked caregiver support strategies.

## Practical Suggestions

This study highlights the need to reframe a diagnosis of dementia not only as a form of clinical disclosure but also as a starting point for caregiver support. Our findings show that providing information about dementia itself and about local medical institutions at the time of diagnosis is significantly associated with a lower caregiver burden.

To translate these findings into practice, standardized protocols should ensure that in addition to the diagnosis, families receive clear explanations of the symptoms and progression of dementia as well as contact points for ongoing medical care in the community. Delivering this information early when families are beginning to adjust can reduce uncertainty, support planning, and promote more stable caregiving at home.

Additionally, physicians involved in diagnosis should be supported through training or resources that help them understand and communicate available local care pathways. Strengthening this role can ensure that caregivers are not left to navigate the system alone.

Positioning the diagnostic moment as a foundation for tailored information delivery can improve continuity of care and reduce long-term stress on family caregivers.

## Limitations

Several limitations should be acknowledged. While this study highlights the importance of types of informational support at the time of dementia diagnosis, we were unable to assess how the information was delivered or the specific content, quality, and source of each type of support. Additionally, because informational support was self-reported, factors such as accuracy, clarity, and the method of communication were not captured objectively. Moreover, responses regarding informational support were based on retrospective recall, as participants reported on their experiences after some time had passed since the diagnosis. This may have introduced recall bias or may have been influenced by subjective interpretation. Finally, the response rate was relatively low, which may have limited the statistical power to detect significant associations. Furthermore, the sample may not fully reflect the diversity of caregiving experiences across Japan. Future research with larger, more representative samples and detailed assessments of informational support, including its timing, format, and delivery context, is needed to deepen understanding and guide the development of effective caregiver support strategies.

## Author Contributions

**TW, TO, TY, and KI** were responsible for the study planning and survey development, data analysis, and manuscript drafting and revision. **TH** assisted the main authors and provided practical guidance for the study’s practical planning. All the authors contributed to the writing and approved the final manuscript.

## Statements and Declarations

### Ethical Considerations

This study was ethically approved by the Tokyo Metropolitan Institute of Gerontology Institutional Review Board, no. R24-084. This study complied with the Declaration of Helsinki and its amendments or comparable ethical standards for conducting surveys.

### Consent to Participate

Participation was considered to indicate consent by returning the completed questionnaire. A checkbox was also provided on the questionnaire for participants who wished to withdraw. Responses returned with this box unchecked were regarded as consent to participate in the study.

### Consent for Publication

Informed consent, which included consent for publication, was obtained from all participants prior to data collection.

### Data Availability

Owing to the sensitive nature of the data collected, the datasets are not publicly available but are available from the corresponding author upon reasonable request and with appropriate ethical approval.

---

**Table 1.** Baseline characteristics (N = 159)

| Characteristics | Total % |
| --- | --- |
| CG gender |  |
| Female | 66.7 |
| Male | 33.3 |
| CG age, mean ± SD | 63.2 ± 11.6 |
| Employment |  |
| Unemployed | 42.1 |
| Employed | 56.0 |
| On leave | 1.9 |
| Financial status |  |
| Very wealthy | 5.7 |
| Wealthy | 21.4 |
| Average | 40.9 |
| Somewhat poor | 23.9 |
| Poor | 8.2 |
| CR gender |  |
| Female | 69.2 |
| Male | 30.8 |
| CR age, mean ± SD | 80.4 ± 8.8 |
| Coresidence |  |
| Live-in | 71.7 |
| Live-out | 28.3 |
| Diagnosis |  |
| Alzheimer | 66.7 |
| Else | 11.9 |
| MCI | 6.9 |
| DK | 14.5 |
| BPSD: DBD5 (0–5), mean ± SD | 1.8 ± 1.4 |
| Time since diagnosis (years), mean ± SD | 4.2 ± 3.6 |
| J-ZBI_8 (0–32), mean ± SD | 13.4 ± 8.2 |

Abbreviations: CG: caregiver; SD: standard deviation; CR: care-recipient; MCI: Mild Cognitive Impairment; DK: don't know; BPSD: Behavioral and Psychological Symptoms in Dementia; DBD: dementia behavior disturbance scale 5-item version; J-ZBI\_8: Japanese version of the Zarit burden inventory.

**Table 2.** Informational support received at the hospital following dementia diagnosis (N = 159)

| Diagnosis type | Total % |
| --- | --- |
| Dementia | 73.6 |
| Long-term care services & community-based integrated care program | 58.5 |
| Legal support, including adult guardianship, abuse prevention, and independent living assistance | 22.6 |
| Financial support systems, including pensions, public assistance, and medical expense subsidies | 22.0 |
| Family caregiver support groups | 32.7 |
| Peer support meetings for persons living with dementia | 25.2 |
| Local medical institutions and clinic | 35.8 |
| Psychological support services for persons with dementia and their family members | 32.1 |
| Employment support for caregivers, including caregiving leave and work-care balance programs | 10.7 |

**Table 3.**
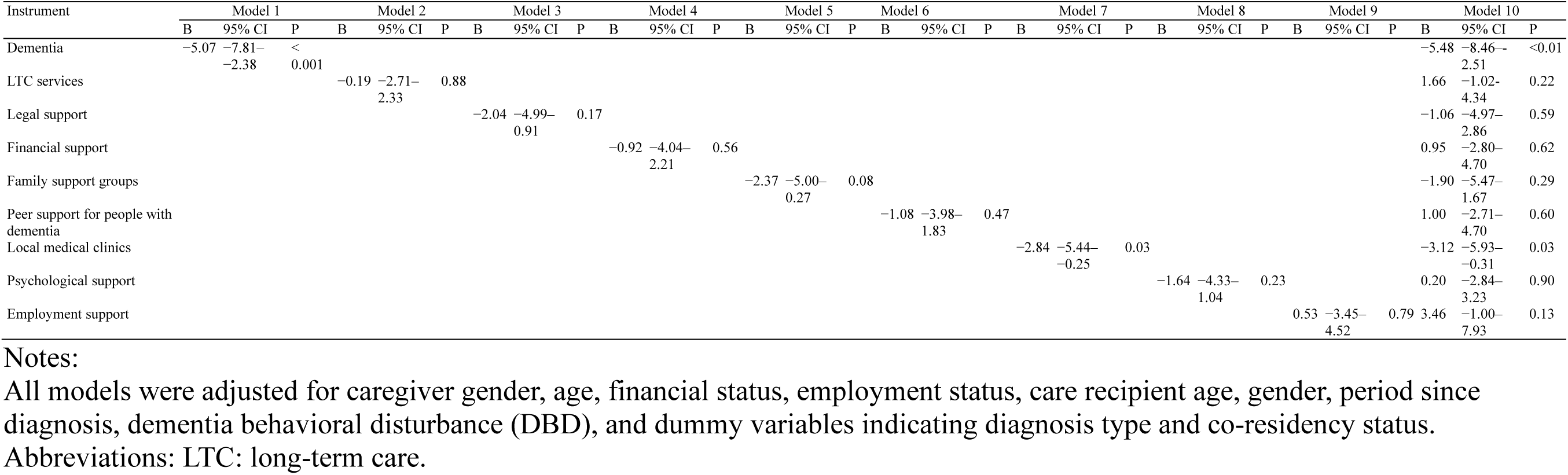
Regression analysis predicting family caregiving burden (n = 159)

**Table 4.** Regression analysis predicting personal and role strain subscales of family caregiving burden (n = 159)

| Instrument | Personal strain |  |  | Role strain |  |  |
| --- | --- | --- | --- | --- | --- | --- |
|  | B | 95% CI | P | B | 95% CI | P |
| Dementia | -3.16 | -5.18–-1.15 | 0.00 | -2.35 | -3.60–-1.10 | 0.00 |
| LTC services | 0.83 | -0.99–2.64 | 0.37 | 0.83 | -0.30–1.95 | 0.15 |
| Legal support | -0.87 | -3.52–1.79 | 0.52 | -0.17 | -1.82–1.48 | 0.84 |
| Financial support | 0.73 | -1.82–3.27 | 0.57 | 0.20 | -1.38–1.78 | 0.80 |
| Family support groups | -0.96 | -3.38–1.45 | 0.43 | -0.92 | -2.43–0.58 | 0.23 |
| Peer support for people with dementia | 0.71 | -1.80–3.22 | 0.58 | 0.32 | -1.24–1.88 | 0.69 |
| Local medical clinics | -2.06 | -3.97–-0.16 | 0.03 | -1.06 | -2.25–0.12 | 0.08 |
| Psychological support | 0.16 | -1.90–2.21 | 0.88 | 0.03 | -1.25–1.31 | 0.96 |
| Instrument | 2.07 | -0.96–5.10 | 0.18 | 1.38 | -0.50–3.26 | 0.15 |

Notes:
Both models were adjusted for caregiver gender, age, financial status, employment status, care recipient age, gender, period since diagnosis, dementia behavioral disturbance (DBD), and dummy variables indicating diagnosis type and co-residency status.
Abbreviations: LTC: long-term care.

